# A rapid and connected test for olfactory loss screening

**DOI:** 10.64898/2026.07.23.26358424

**Authors:** E Moussy, A Fournel, C Daudé, M Fieux, C Ferdenzi, M Bensafi, M Richard

**Affiliations:** Université Lyon 1, CNRS, INSERM, CRNL, UMR 5292, UMR S1028, Bron, France; Laboratoire d’Étude des Mécanismes Cognitifs (EA3082), Université Lyon 2, 69676 Bron, France; Hospices Civils de Lyon, Centre Hospitalier Lyon Sud, Service d’ORL, d’otoneurochirurgie et de chirurgie cervico-faciale, Pierre Bénite cedex F-69310, France; Université de Lyon, Université Lyon 1, F-69003, Lyon, France

**Keywords:** Olfactory evaluation, smell, psychophysical test, screening, olfactory disorders, anosmia

## Abstract

**INTRODUCTION:** Despite their widespread prevalence and deleterious consequences, olfactory disorders remain little evaluated in clinical practice and neuroscientific studies, due to the lack of early, quick and reliable detection tools. To meet this demand, we developed a new connected olfactory test and evaluated its ability to discriminate dysosmia (i.e., altered sense of smell) from normosmia.

**METHODS:** The Olfactory Screening with Connected And Rapid Detection test (OS-card, version 1.0) consisted of 8 RFID-equipped scratch-and-sniff cards interfaced with a dedicated mobile application. In this test, each micro-encapsulated odor is rated for perceived intensity on a 5-point scale and identified using a 4-alternative forced-choice procedure. The olfactory abilities of two matched samples of 25 normosmic and 25 dysosmic participants were evaluated using the OS-card 1.0 and two reference tests: the European Test for Olfactory Capabilities ETOC and the identification subtest of the Sniffin’ Sticks.

**RESULTS:** Intensity and identification OS-card scores were significantly lower in dysosmics than in normosmics. OS-card was easy to use and took less than 10 minutes to complete. OS-card scores were well correlated with ETOC scores and Sniffin’ Sticks scores. According to a cross-validation approach using olfactory status categories provided by the ETOC, it was found that OS-card correctly classified 100% of the normosmic participants, 68% of the dysosmics (hyposmics and anosmics together), and 96% of the anosmics.

**CONCLUSION:** OS-card is user friendly, rapid and reliable for detecting severe smell disorders (anosmia).

## Introduction

The sense of smell plays an important, though long underestimated, role in humans’ everyday life – especially in food intake, social interactions and danger detection. Consequently, losing it has harmful consequences on living conditions ^(1)^. Olfactory disorders range from quantitative deficits including hyposmia (a reduced olfactory perception) and anosmia (a complete loss of olfaction) to qualitative modifications (parosmia and phantosmia) ^(2)^. Their prevalence is not negligible since it reaches around 15-20% in Europe and the USA ^(3–6)^. These estimates date from before the Covid-19 pandemic, which undoubtedly exacerbated these figures (due to persistent olfactory losses following COVID-19 infection: e.g. ^(7–11)^).

Despite their prevalence and impact in the population, olfactory disorders are poorly managed in terms of diagnosis. Ideally, anyone with suspected olfactory disorders may initiate medical consultation, that will infirm or confirm these suspicions and start appropriate care if needed. However, this process is not optimal, mainly because of the lack of suitable tools to evaluate olfactory abilities. Firstly, using an objective evaluation is essential since the diagnostic based on self-assessment of one’s own olfactory status has repeatedly proven to be unreliable ^(4,12,13)^. Secondly, although robust tools for psychophysical testing exist, such as the UPSIT (University of Pennsylvania Smell Identification Test) ^(14)^, the Sniffin’ Sticks ^(15)^ and the ETOC (European Test of Olfactory Capability) ^(16)^, they are rarely used in routine medical consultations ^(17)^. Taste and Smell Clinics of course have these psychophysical tests available, but they are few and not easily accessible to the patients as they are not financially covered ^(18)^. The reason why psychophysical testing is not widely used in the Ear Nose and Throat (ENT) consultations ^(19)^, and even less so in general consultations, is that these tests are difficult to implement in everyday practice. The main disadvantage of the reference tests mentioned above is that they take too long, in addition to being restrictive in terms of storage and preservation. Furthermore, they are not single-use, which can be problematic in case of contagious diseases, and they are not designed for self-administration, which limits their application for screening. It should be noted that these issues also apply to basic research, where many neuroscientific studies continue to use self-assessment to evaluate the olfactory function of their volunteer participants.

The limited use of objective olfactory evaluation tools in medical consultations both leads to a probable underestimation of the prevalence of olfactory disorders and contributes to the patients’ dissatisfaction with the way they are cared for. In 2009, 60% of Swiss and German patients attending a consultation in a Smell and Taste Clinic reported being not or poorly informed about their olfactory disorder and its potential consequences ^(20)^. According to a more recent survey in France, almost all patients (94%) attending an ENT consultation or members of a patient association were dissatisfied with the way their olfactory disorder was managed ^(21)^. This concerning trend highlights the growing need for healthcare professionals to address the Covid-19-related increasing awareness of olfactory disorders and their consequences. Thus, in order to improve access to care for patients with dysosmia, we have developed a new psychophysical olfactory test, The Olfactory Screening with Connected And Rapid Detection test (OS-card, version 1.0). Here, we present this test, which was designed to be rapid, single-use, self-administered and user-friendly. OS-card ability to screen for olfactory disorders was tested by comparison with two reference tests, ETOC and the Sniffin’Sticks.

## Material and method

### Participants

A total of 50 French adult participants (25 normosmics and 25 dysosmics, as classified by the ETOC test ^(16)^) were recruited from January 2024 to June 2024 (see **Supplementary Material and Table S1** for demographic information and **Supplementary Material** for ETOC description). Participants were recruited through different distribution channels such as associations, open university, community center for social action, volunteers’ database, hospitals, or by word of mouth. Dysosmic participants, including 14 hyposmics and 11 anosmics, covered a variety of etiologies (3 post-Covid-19, 3 post-traumatic, 3 chronic rhinosinusitis with or without polyposis, 1 polyposis with rhinosinusitis, 1 post-surgical tumoral intervention and 14 idiopathics probably including cases of age-related loss of smell, known as presbyosmia). Informed written consent was obtained from all participants. This study was performed in accordance with the Declaration of Helsinki on Biomedical Studies involving human subjects. The study design and participant consent procedure were approved by the South-Est III ethic review board (2022) (RCB ID number: 2022-A01042-41).

### Procedure

The olfactory performances of the participants were evaluated during a single session, using three different psychophysical tests: two standard psychophysiological tests, the ETOC test ^(16)^ and the Sniffin’ Sticks identification subtest ^(15)^, and the newly developed OS-card 1.0. The olfactory tests were administered in that order, with 4-min breaks between the tests.

### OS-card olfactory test

OS-card 1.0 comprises 8 scented cards equipped with an RFID (Radio Frequency IDentification) chip (adhesive NFC (Near Field Contact) tags; Circus NTAG210) (**Figure 1A**), an NFC reader (ACR1255U-J1; Advanced Card Systems) (**Figure 1B**) and the software Inspir-O developed in house and installed on an Android tablet (Lenovo Tab M10 Plus) (**Figure 1C**). Eight odors mainly from the food sphere, were included (banana, cinnamon, clove, eucalyptus, garlic, mint, orange, thyme, see **Supplementary Material and Table S2** for a description of the odorants and of the odor selection process).

**Figure 1.**
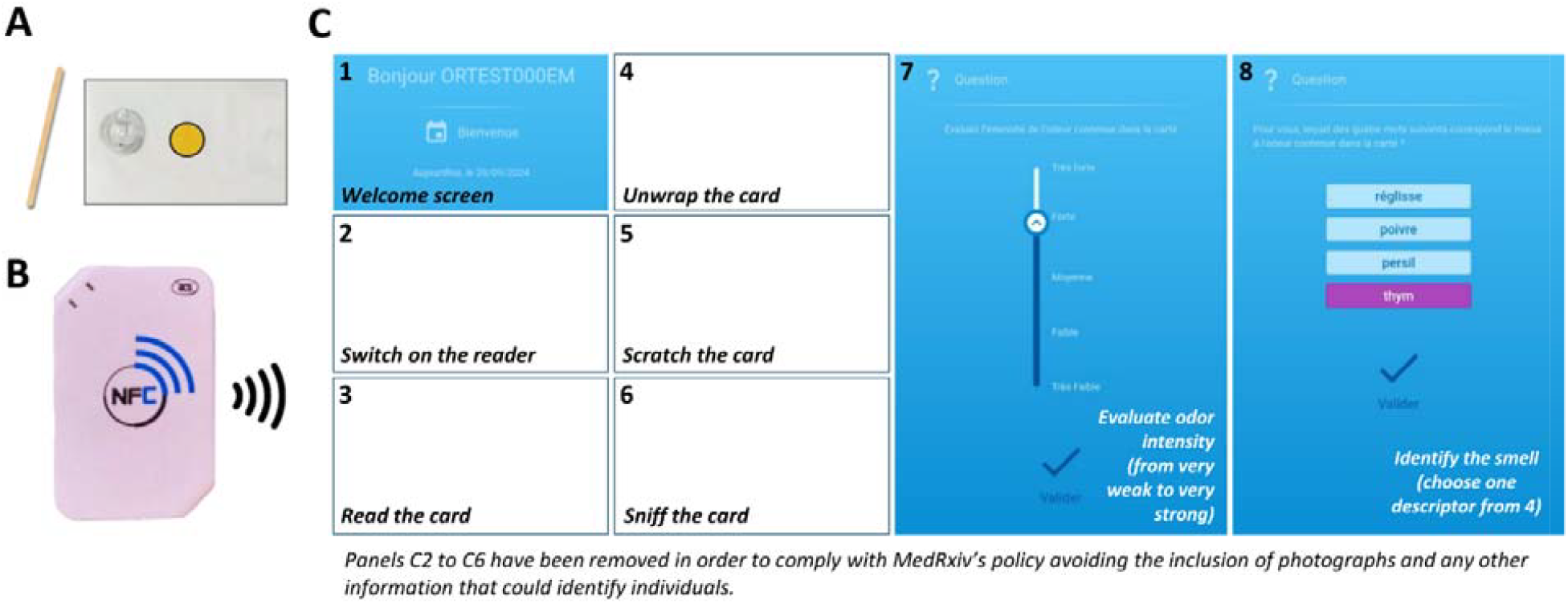
OS-card test material. A) An odorized card with its wooden stick. B) The Bluetooth NFC reader. C) Screenshots of the instructions given via the digital tablet. Instructions were provided in French and translated here for the purpose of the publication.

The test was conducted as follows. At each step of the procedure, the software assisted in detail the participants with video instructions displayed via the digital tablet, allowing them to perform the test in autonomy. The cards were numbered from 1 to 8, indicating the order in which they were to be smelled, and were presented in the same order to all the participants: 1-clove, 2-orange, 3-thyme, 4-cinnamon, 5-eucalyptus, 6-banana, 7-garlic, 8-mint. In this study, an experimenter was always present to respond to questions if necessary. Participants were instructed to:

1. read general instructions via a welcome screen (**Figure 1C-1**),
2. switch ON the NFC reader (**Figure 1C-2**),
3. read the first card via the Bluetooth NFC reader (**Figure 1C-3**),
4. unpack the card (**Figure 1C-4**),
5. scratch the yellow-colored circle of the card using a clean wooden stick (**Figure 1C-5**),
6. sniff the card (**Figure 1C-6**),
7. rate the intensity of the odor (very weak, weak, medium, strong or very strong) (**Figure 1C-7**),
8. identify the odor among four proposed items (**Figure 1C-8**).
9. repeat the procedure for each of the seven other cards.

Participants were allowed to sniff the card as many times as necessary before responding.

### Reference olfactory tests

**The ETOC** ^(16)^ **and the Sniffin**’ **Sticks identification subtest** ^(15)^, two reference olfactory tests were used as described in previous publications. The ETOC yielded a detection/localization score (between 0 and 16) and an identification score (between 0 and 16). Each participant was classified as normosmic (no olfactory disorde ), hyposmic (reduced olfactory perception) or anosmic (complete or almost complete loss of olfaction), using these two scores and according to normative data from a previous study ^(22)^. The Sniffin’ Sticks yielded an identification score (between 0 and 16). A detailed description of each test is provided in **Supplementary Material**.

### Data analysis

All statistics were performed using R software ^(23)^ and alpha value was set to 0.05. To evaluate OS-card test performance, normosmic and dysosmic participants (according to ETOC) were compared using Wilcoxon or Chi-Square tests across four parameters: per-card intensity ratings and correct identification rates, as well as each participant’s mean intensity score and overall identification score. Correlation analyses between the OS-card scores and those of the reference olfactory tests were performed using Spearman correlations. Finally, to evaluate the ability of OS-card to accurately discriminate between normosmic and dysosmic participants, we used linear models on which we performed a discrimination analysis using cross-validation (with a leave one out cross-validation) using the package ‘Caret’ ^(24)^. Classification performances were assessed using the Area Under the Curve (AUC) of the mean Receiver Operating Characteristic (ROC), the total correct classification rate (rate of correctly classified participants), the sensitivity (true positive rate, i.e. rate of correctly classified dysosmics) and the specificity (false positive rate, i.e. rate of correctly classified normosmics). We used a criterion of probability equal to or higher than 50% to affect a participant to a specific group (normosmics or dysosmics). Three different models were tested: *normosmics vs dysosmics* using i) both intensity and identification OS-card scores or ii) only identification, and *normosmics vs anosmics* using both scores. See more details in **Supplementary Material**.

## Results

### OS-card test performances

The perceived intensity was significantly higher in normosmics than in dysosmics for 5 odors (**Figure 2A**): banana, cinnamon, clove, eucalyptus and garlic (Wilcoxon test with Bonferroni correction: W=145.50, p<0.01; W=166, p=0.045; W=118.50, p<0.001; W=153.50, p=0.01; W=148, p<0.01, respectively). Similar trends were observed for mint, orange and thyme (W=178.5, p=0.056; W=186.5, p=0.08; W=179, p=0.054, respectively). On average, the mean intensity score was higher for normosmics (3.38±0.10) than for dysosmics (2.30±0.22; W = 500, p <0.001) (**Figure 2B**).

**Figure 2.**
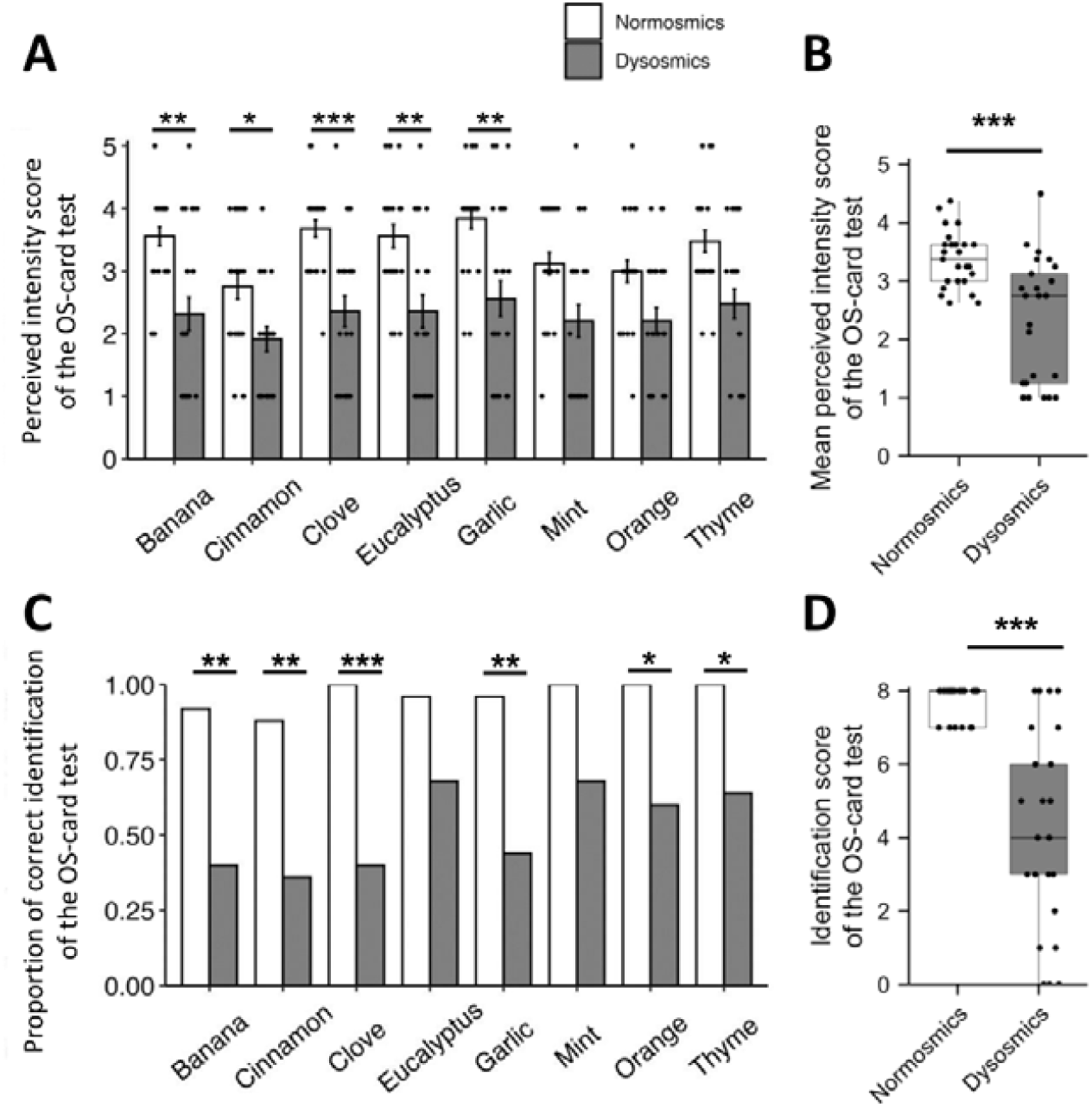
OS-card scores in participants classified as normosmic (n=25) and dysosmic (n=25) according to ETOC. **A)** Perceived intensity score for each scented card (mean±sem). **B)** Mean perceived intensity score. **C)** Proportion of correct identification for each scented card. **D)** Identification scores. On the boxplot representation (B and D), the outer box range represents the first and third quartiles, the middle horizontal black line represents the median. Vertical lines join data points that are included between the first quartile – 1.5 IQR and third quartile + 1.5 IQR. Dots represent individuals. **\* p<0.05, ** p<0.01, *** p<0.001**.

Regarding odor identification, the proportion of correct classification was higher in normosmics than in dysosmics for 6 odors: banana, cinnamon, clove, garlic, orange and thyme (Chi-Square test with Bonferroni correction: *X* ^*2*^_*(1)*_=12.83, p<0.01; *X* ^*2*^_*(1)*_=12.22, p<0.01; *X* ^*2*^_*(1)*_=18.67, p<0.001; *X* ^*2*^_*(1)*_=13.71, p<0.01; *X* ^*2*^_*(1)*_=10.13, p=0.012; *X*^*2*^ _*(1)*_=8.67, p=0.026 respectively). The difference did not reach significance for eucalyptus and mint (*X* ^*2*^_*(1)*_=4.88, p=0.22 ; *X*^*2*^ _*(1)*_=7.29, p=0.055 respectively) (**Figure 2C**). In accordance with these results, the identification score was significantly higher in normosmics (7.72±0.09) than in dysosmics (4.20±0.53; W = 554, p <0.001) (Figure 2D).

Taking the OS-card test in autonomy took less than 10 minutes in total.

### Correlations between OS-card and reference tests scores

Correlation analyses showed that the OS-card perceived intensity score was positively correlated with the localization score of the ETOC test (Spearman test: S=8434, r_s(48)_=0.60, p<0.0001) (**Figure 3A**). For the identification scores, we found positive correlations between OS-card and ETOC (S=3673.7, r_s(48)_=0.82, p<0.0001) (**Figure 3B**), and between OS-card and Sniffin’ Sticks (S=4129.10, r_s(48)_=0.80, p<0.001) (**Figure 3C**).

**Figure 3.**
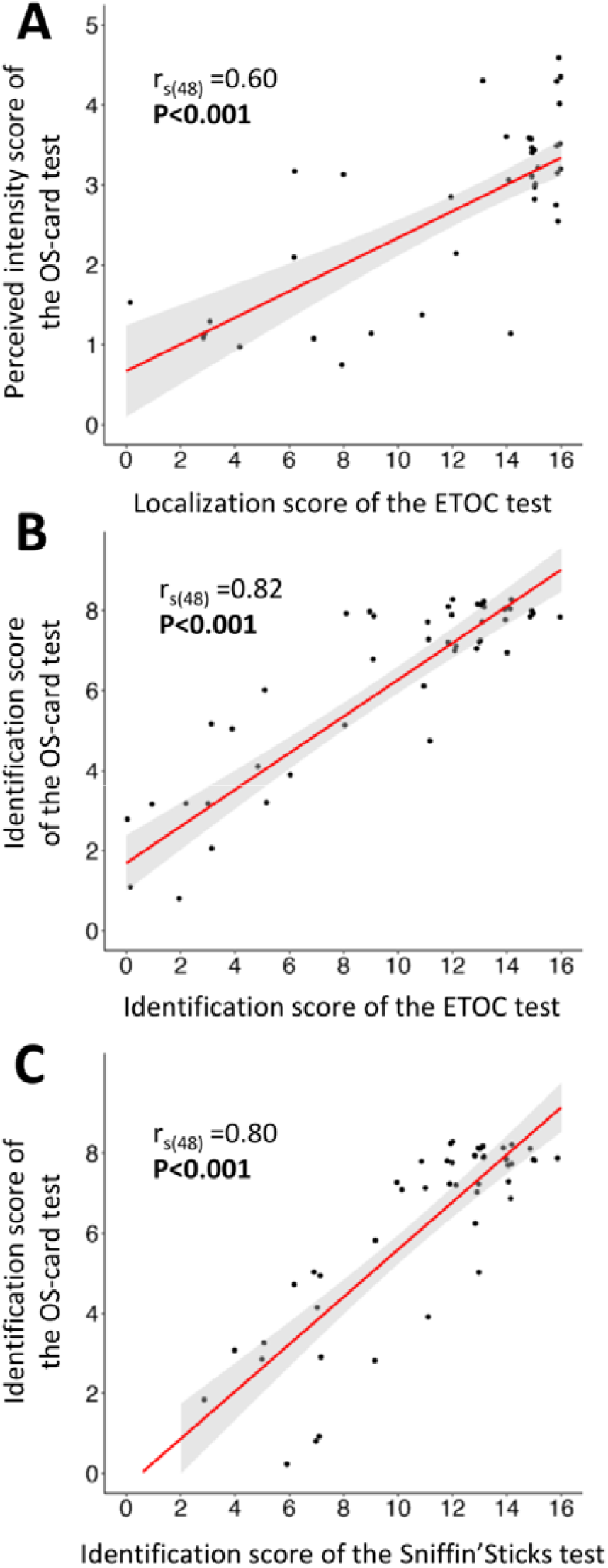
Correlations between olfactory scores obtained with OS-card and those obtained with the reference tests ETOC and Sniffin’ Sticks. **A**) Positive correlation between the OS-card intensity score and the ETOC localization score. **B**) Positive correlation between the OS-card and the ETOC identification scores. **C**) Positive correlation between the OS-card and the Sniffin’ Sticks identification scores. A linear smooth line is represented in red. Dots represent individuals.

### Discrimination analysis

#### SNormosmics vs dysosmics

To test to what extent the OS-card identification and intensity scores allow to accurately discriminate normosmics from dysosmics, we ran a first linear discrimination model. Misclassifications were only false negatives due to 8 dysosmics classified as normosmics (**Figure 4A**), for a total correct classification rate of 84% (42 out of 50 participants), a sensitivity of 68% (17 dysosmics correctly classified out of 25) and a specificity of 100% (25 correctly classified normosmics out of 25). Area Under the Curve (AUC) of the mean Receiver Operating Characteristic (ROC) was 0.82 (**Figure 4B**). The probabilities of being dysosmic according to the combination of the two OS-card scores are depicted in **Figure 4C**. The 8 misclassified dysosmics were all borderline hyposmics, who had higher OS-card intensity score (3.36±0.20) and identification score (7.25±0.31) than the correctly classified dysosmics (intensity 1.80±0.21, W=11, p<0.001; identification: 2.77±0.44, W=0, p<0.001). These misclassified dysosmics were al o significantly older (69.25±1.60 versus 59.94±3.22 years in the correctly classified dysosmics: t_(22)_=-2.59, p=0.02). Note that the model used primarily the identification score (linear discrimination coefficient: -1.46) rather than the intensity score (linear discrimination coefficient: 0.11) to discriminate between groups.

**Figure 4.**
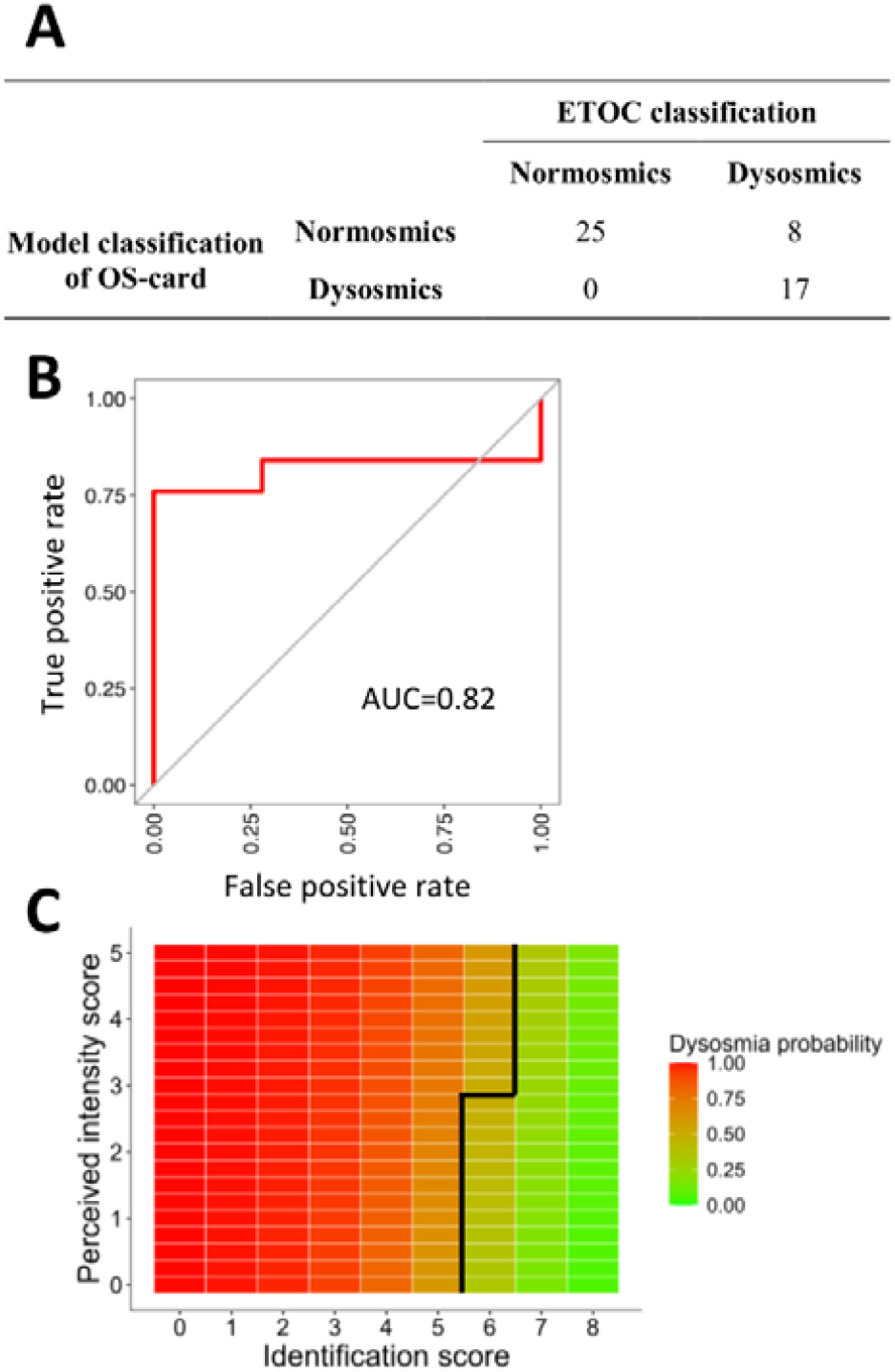
Probability of dysosmia according to OS-card. **A**) Confusion matrix of the discrimination analysis (normosmics vs dysosmics). **B**) Mean ROC curve of the 49 acquired models after the leave-one-out cross-validation. The curve represents the true positive rate (sensitivity) for the false positive rate (specificity). **C**) Probability of being dysosmic according to the OS-card intens ty and identification scores. The black line represents the cut-off for which the combination of the two scores is associated with a probability of normosmia >50%.

For this reason, and also because other tests perform reliable classification based solely on identification, we ran a second model using only the OS-card identification score. The results were identical to the first model that included both scores, with an AUC of the mean ROC equal to 0.82 and a total correct classification rate of 84% (sensitivity=68%, specificity=100%). The 8 misclassified dysosmics were the same as in the initial model that included both scores. The probabilities of being normosmic or dysosmic according to the OS-card identification score alone are provided in **Table 1**. Participants with an OS-card identification score of 6 or more are classified as normosmics, and those with a lower score as dysosmics.

**Table 1.** Probability of normosmia and dysosmia according to the classification model using only the identification score.

| Identification score | Normosmia probability | Dysosmia probability |
| --- | --- | --- |
| 0 | 0.003 | 0.997 |
| 1 | 0.008 | 0.992 |
| 2 | 0.021 | 0.980 |
| 3 | 0.052 | 0.947 |
| 4 | 0.129 | 0.871 |
| 5 | 0.281 | 0.719 |
| 6 | 0.510 | 0.490 |
| 7 | 0.734 | 0.266 |
| 8 | 0.880 | 0.120 |

#### Normosmics vs anosmics

Last, to test the ability of OS-card to discriminate between more extreme olfactory statuses, we ran a third model (using both intensity and identification scores) comparing normosmics and anosmics. Only one anosmic individual was misclassified (traumatic etiology; intensity score: 2.13; identification score: 5), leading to near perfect classification performances (**Figure 5A**): AUC of the mean ROC = 1 (**Figure 5B**), total correct classification rate = 97.2% (35 out of 36 participants), sensitivity = 91% (10 anosmics correctly classified out of 11), specificity = 100% (25 correctly classified normosmics out of 25). The probabilities of being anosmic according to the combination of the two OS-card scores are depicted in **Figure 5C**.

**Figure 5.**
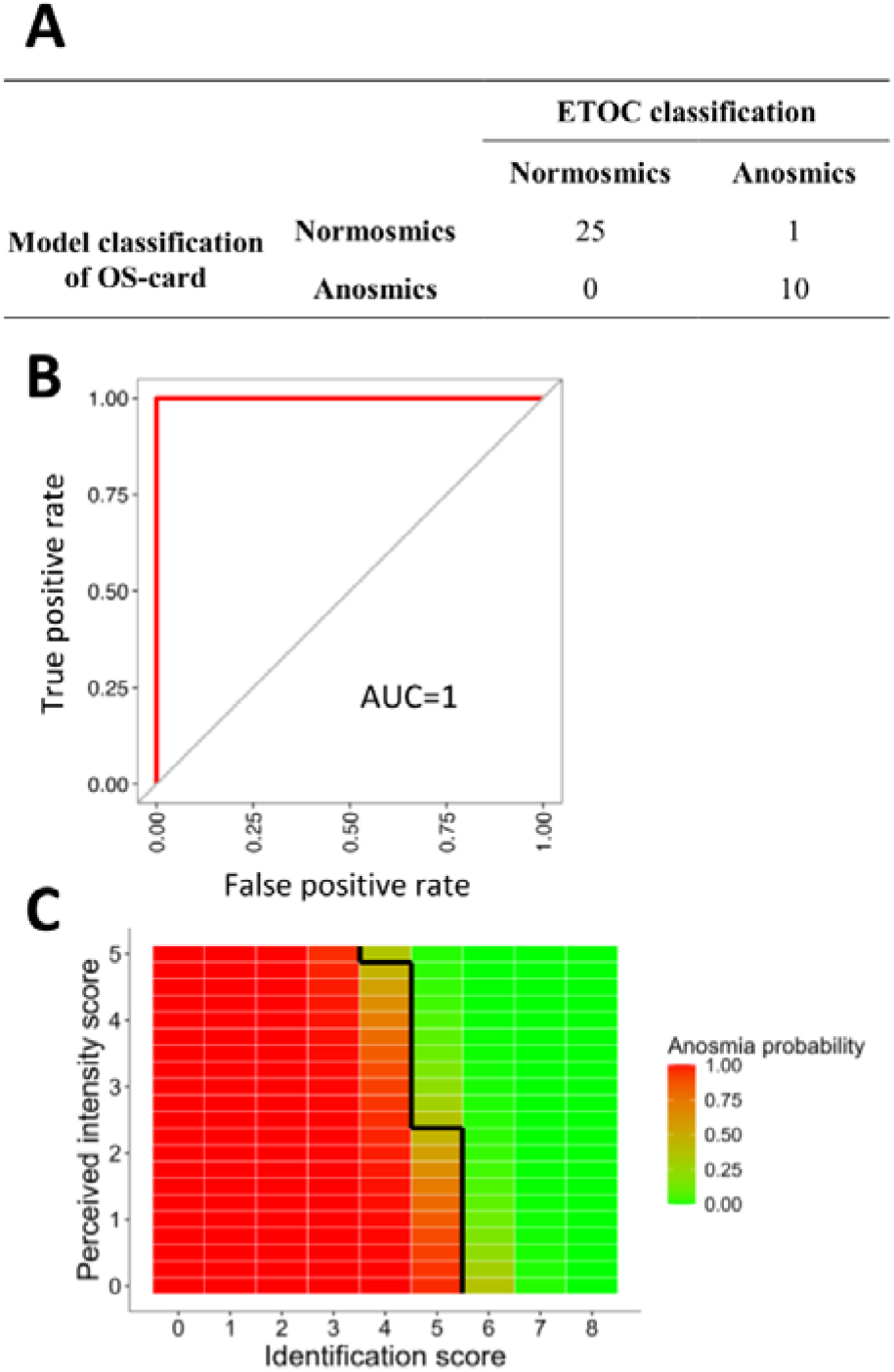
Probability of anosmia according to OS-card. **A**) Confusion matrix of the discrimination model (normosmics vs anosmics). **B**) Mean ROC curve of the 35 acquired models after the leave-one-out cross-validation using a down-sampling method. The curve represents the true positive rate (sensitivity) for the false positive rate (specificity). B) Probability of being anosmic according to the OS-card intensity and identification scores. The black line represents the cut-off for which the combination of the two scores is associated with a probability of normosmia >50%.

## Discussion

The aim of the present study was to develop a reliable, rapid and user-friendly olfactory test to screen for olfactory disorders. OS-card 1.0 is the first version of a test based on 8 single-use scratch-and-sniff cards connected to a phone application collecting odor intensity ratings and identification responses. Using it in 25 normosmic and 25 dysosmic participants, who also took two reference olfactory tests (ETOC and Sniffin’ Sticks), revealed that OS-card is a reliable test for detecting severe smell disorders (anosmia), while being quick (less than 10 minutes) and easy to use for self-administration.

In fact, we first found that OS-card 1.0 has good performances. The perceived intensity ratings and the olfactory identification scores obtained with OS-card were significantly lower in people with a loss of smell than in control participants with no olfactory loss. In addition, OS-card scores were highly consistent with those of more comprehensive tests of reference, as shown by the high positive correlations with ETOC scores ^(16)^ and Sniffin’ Sticks scores ^(15)^, especially for odor identification (r_s(48)_>0.80). This performance is high in itself, but also in comparison to other newly developed olfactory tests based on odor identification (r=0.74 in ^(25)^; r=0.52 in ^(26)^; although the latter used a threshold test – measuring a different olfactory ability – as comparator).

Regarding the ability of OS-card 1.0 to categorize people as having an olfactory impairment or not, we found near-perfect ability to identify severe olfactory loss. Indeed, when comparing extreme groups (*normosmics vs anosmics* according to ETOC) with OS-card, we found 100% specificity (all normosmics categorized as such) and 91% sensitivity (only one anosmic individual misclassified as normosmic). When the whole range of olfactory impairment is considered, including also mild olfactory impairment (*normosmics* vs *dysosmics*), specificity remained perfect with 100% of normosmics classified as such, but sensitivity was lower with 68% of dysosmics correctly classified. The same levels of specificity and sensitivity were found when, instead of taking both intensity and identification scores into account, we included only identification responses. The misclassified dysosmic individuals did not have a pronounced olfactory disorder: The fact that they were borderline to their category made it difficult for the classification model to differentiate them from normosmics with low scores. They also were 10 years older than the correctly classified dysosmic participants, suggesting that age-related loss of smell ^(27)^ may be more difficult to pinpoint with OS-card 1.0. This observation may also explain why sensitivity was a little lower here compared to a preliminary version of OS-card used in younger subjects (although it included fewer scented cards: 4 cards, 70% sensitivity) ^(28)^. However, it should be noted that overall, the correct classification rate remains higher in the 8-card OS-card test than in the 4-card test (84% in the present study, versus 73% in ^(28)^).

In addition to these considerations related to the reliability of the OS-card 1.0, other major advantages of this test are worth highlighting: Its ease of use, short duration, and the possibility of doing it yourself. Several brief tests have been proposed for olfactory screening ^(29–32)^, and remote testing has been implemented in some ^(33,34)^. Our test is thus no exception, but it has the advantage of working with a phone and tablet app, an option that is not very common, even though we have seen several recent examples of digital support ^(25,26,34,35)^. This connected support includes detailed steps for using the test, combined with intuitive and fun operation of the olfactory cards, making the self-administration smooth and enjoyable. Moreover, the OS-card 1.0 was quick since it took less than 10 minutes to complete (instructions included). Taken together, these features suggest that OS-card may be suitable for clinical and research contexts in which time constraints are a key consideration.

With a view to improving OS-card performance for discriminating normosmics from dysosmics (which was less effective than the normosmics vs anosmics categorization), a few methodological choices can be discussed. Among the two parameters used for olfactory status classification, intensity did not have a significant impact, since removing it did not alter the performances of the model. Also, intensity was counterintuitively used by the classification model, since participants with identification scores of 6 were classified as normosmics if they rated intensity below 3 but dysosmics above 3 (*Figure 4*). While intensity is an important olfactory dimension in our and other studies ^(34)^ for discriminating normosmia from anosmia, this task may be less well suited for more subtle discrimination. Intensity rating is undoubtedly less objective than identification (forced choice among four possibilities) since it may depend on how each person uses the rating scale. For example, it may be that borderline normosmics and borderline hyposmics used the intensity scale differently, triggering the surprising classification mentioned above. In this respect, a “blank” card with no odor (see ^(28)^) would have been useful to provide a common reference and to try to reduce subjectivity in the scale use. Alternatively, one may consider the opportunity of using only identification to categorize the participants as normosmic or dysosmic, as is the case in other olfactory tests ^(36)^. With OS-card, the threshold of predicted normosmia >50% corresponded to at least 6/8 correct identifications, which is consistent with the threshold of reference tests (around 12/16 for the ETOC identification test and 11/16 for the Sniffin’ Sticks) ^(15,16)^.

Finally, several technical improvements could be implemented in future versions of OS-card. For instance, we chose to individually wrap the cards in a plastic packaging to protect them, and to use sticks to release each scent from the microcapsules. These two technical choices can be perfectly replaced by a more convenient solution, such as a detachable transparent sticker placed on the microcapsules. This is used for example in the recently developed SCENTinel screening test ^(34)^: It releases the smell when the sticker is detached and preserves the olfactory stimulus while limiting the ecological impact of the test.

In conclusion, based on the results of this study, our recommendations for future users are as follows. To determine if a patient suffers from anosmia, OS-card is a reliable and quick olfactory test. Screening for severe olfactory loss is thus possible with the test as it is presented in this article. Alternatively, to get an indication of the presence of an olfactory loss, without necessarily defining it very precisely (i.e., as anosmia or hyposmia), then using OS-card identification score alone may be sufficient. However, it must be kept in mind that the classification is rough (this is the inevitable disadvantage of rapid tests), especially in the intermediate scores. In this case, further examination using a more complete olfactory evaluation tool may be considered (e.g., with the ETOC ^(16)^), especially if the OS-card outcome is “normosmic” despite a significant complaint of the patient.

In terms of applications and perspectives, the OS-card screening test can be aimed at a wide range of audiences. In addition to its usefulness for systematic olfactory screening of participants in neuroscientific and/or psychological studies, OS-card primary function is to be used during clinical assessment. This may concern ENT consultations (which will probably prefer to equip themselves with more accurate assessment tools though, such as ETOC or Sniffin’ Sticks) but also – and maybe more likely – general medical consultations or even pharmacies. The widespread availability of NFC technology on most smartphones ^(37)^, combined with OS-card’s user-friendly self-administration design, paves the way for large-scale, population-wide screening of olfactory function. One of the specific benefits of such a screening would be to track olfactory disorders at an epidemiological level, which could prove particularly useful in a pandemic context ^(9)^. A further area of potential application concerns older adults, who are frequently affected by olfactory impairments that can compromise their well-being and quality of life. More broadly, the widespread use of such sensory screening could help raise public awareness of the importance of the sense of smell in everyday life ^(38)^. Finally, the connected olfactory monitoring system developed for OS-card could also benefit the course of treatment following the detection of an olfactory disorder. Namely, it could be used in the future to develop remote olfactory training programs that allow therapists to monitor patient progress, promote consistent practice, and ultimately improve treatment outcomes.

## Data Availability

The data that has been used is confidential

## References

1. Croy I, Nordin S, Hummel T. Olfactory disorders and quality of life--an updated review. Chem Senses. 2014;39(3):185–94.

2. Hummel T, Nordin S. Olfactory disorders and their consequences for quality of life. Acta Otolaryngol. 2005;125(2):116–21.

3. Brämerson A, Johansson L, Ek L, Nordin S, Bende M. Prevalence of olfactory dysfunction: the skövde population-based study. Laryngoscope. 2004;114(4):733–7.

4. Manesse C, Ferdenzi C, Mantel M, et al. The prevalence of olfactory deficits and their effects on eating behavior from childhood to old age: A large-scale study in the French population. Food Quality and Preference. 2021;93:104273.

5. Mullol J, Alobid I, Mariño-Sánchez F, et al. Furthering the understanding of olfaction, prevalence of loss of smell and risk factors: a population-based survey (OLFACAT study). BMJ Open. 2012;2(6):e001256.

6. Murphy C, Schubert CR, Cruickshanks KJ, Klein BEK, Klein R, Nondahl DM. Prevalence of olfactory impairment in older adults. JAMA. 2002;288(18):2307–12.

7. Boscolo-Rizzo P, Fabbris C, Polesel J, et al. Two-Year Prevalence and Recovery Rate of Altered Sense of Smell or Taste in Patients With Mildly Symptomatic COVID-19. JAMA Otolaryngol Head Neck Surg. 2022;148(9):889–91.

8. Ferdenzi C, Bousquet C, Aguera P-E, et al. Recovery From COVID-19-Related Olfactory Disorders and Quality of Life: Insights From an Observational Online Study. Chem Senses. 2021;46:bjab028.

9. Pierron D, Pereda-Loth V, Mantel M, et al. Smell and taste changes are early indicators of the COVID-19 pandemic and political decision effectiveness. Nat Commun. 2020;11(1):5152.

10. Saccardo T, Roccuzzo G, Fontana A, et al. Long-term self-reported symptoms and psychophysical tests in COVID-19 subjects experiencing persistent olfactory dysfunction: a 4-year follow-up study. Front Neural Circuits [Internet]. 2025 [cited 2026 Apr 22];19. Available from: https://www.frontiersin.org/journals/neural-circuits/articles/10.3389/fncir.2025.1538821/full

11. Stanley HB, Pereda-Campos V, Mantel M, et al. Identification of the needs of individuals affected by COVID-19. Commun Med (Lond). 2024;4(1):83.

12. Landis BN, Hummel T, Hugentobler M, Giger R, Lacroix JS. Ratings of overall olfactory function. Chem Senses. 2003;28(8):691–4.

13. Oleszkiewicz A, Hummel T. Whose nose does not know? Demographical characterization of people unaware of anosmia. Eur Arch Otorhinolaryngol. 2019;276(6):1849–52.

14. Doty RL, Shaman P, Kimmelman CP, Dann MS. University of pennsylvania smell identification test: A rapid quantitative olfactory function test for the clinic. Laryngoscope. 1984;94(2):176–8.

15. Hummel T, Sekinger B, Wolf SR, Pauli E, Kobal G. ‘Sniffin’ Sticks’: Olfactory Performance Assessed by the Combined Testing of Odour Identification, Odor Discrimination and Olfactory Threshold. Chem Senses. 1997;22(1):39–52.

16. Thomas-Danguin T, Rouby C, Sicard G, et al. Development of the ETOC: a European test of olfactory capabilities. Rhinology. 2003;41(3):142–51.

17. Ferdenzi C, Bellil D, Boudrahem S, et al. La rééducation olfactive : bénéfices d’une prise en soins pluri-professionnelle. La Presse Médicale Formation. 2022;3(1, Part 1):5–12.

18. Naimi BR, Hunter SR, Boateng K, et al. Patient Insights into the Diagnosis of Smell and Taste Disorders in the United States. medRxiv. 2023;2023.09.20.23295861.

19. Vandersteen C, Dubrulle C, Manera V, Castillo L, Payne M, Gros A. Persistent post-COVID-19 dysosmia: Practices survey of members of the French National Union of Otorhinolaryngology-Head and Neck Surgery Specialists. CROSS analysis. Eur Ann Otorhinolaryngol Head Neck Dis. 2023;S1879-7296(23)00052-2.

20. Landis BN, Stow NW, Lacroix J-S, Hugentobler M, Hummel T. Olfactory disorders: The patients’ view. Rhinology. 2009;47(4):454–9.

21. Tholin L, Rumeau C, Jankowski R, Gallet P, Wen Hsieh J, Nguyen DT. Experience of French patients with olfactory disorders. European Annals of Otorhinolaryngology, Head and Neck Diseases [Internet]. 2024 [cited 2024 Apr 25]; Available from: https://www.sciencedirect.com/science/article/pii/S1879729624000267

22. Joussain P, Bessy M, Faure F, et al. Application of the European Test of Olfactory Capabilities in patients with olfactory impairment. Eur Arch Otorhinolaryngol. 2016;273(2):381–90.

23. R Core Team. R: A Language and Environment for Statistical Computing. 2023; Available from: https://www.R-project.org/

24. Kuhn M. Building Predictive Models in R Using the caret Package. Journal of Statistical Software. 2008;28:1–26.

25. Bernard BJ, Baker O, Pauley A, et al. A Novel Application-Based Test for Rapid Screening of Olfactory Dysfunction. JAMA otolaryngology--head & neck surgery. 2026;152(3):294–300.

26. Alves de Sousa F, Nakanishi M, Santos M, Nóbrega Pinto A, Galvão C, Hummel T. Digitizing Olfactory Assessment in Portugal: Pilot Clinical Application of a Digital Odor Identification Test. ORL. 2025;1–10.

27. Doty RL, Kamath V. The influences of age on olfaction: a review. Front Psychol. 2014;5:20.

28. Moussy E, Fournel A, Bellil D, et al. Losing olfaction in COVID-19: Screening, training and effects on quality of life. Clinical Nutrition Open Science. 2024;56:49–64.

29. Jackman AH, Doty RL. Utility of a three-item smell identification test in detecting olfactory dysfunction. Laryngoscope. 2005;115(12):2209–12.

30. Mueller C, Renner B. A new procedure for the short screening of olfactory function using five items from the “Sniffin’ Sticks” identification test kit. Am J Rhinol. 2006;20(1):113–6.

31. Said M, Davis P, Davis S, Smart K, Davis S, Yan CH. A Rapid Olfactory Test as a Potential Screening Tool for COVID-19. JAMA Otolaryngol Head Neck Surg. 2021;147(9):828–31.

32. Shiga H, Yamamoto J, Kitamura M, et al. Combinations of two odorants of smell identification test for screening of olfactory impairment. Auris Nasus Larynx. 2014;41(6):523–7.

33. Jaén C, Maute C, Mackin S, et al. Remote olfactory assessment using the NIH Toolbox Odor Identification test and the brain health registry. PLoS One. 2024;19(4):e0301264.

34. Parma V, Hannum ME, O’Leary M, et al. SCENTinel 1.0: Development of a Rapid Test to Screen for Smell Loss. Chem Senses. 2021;46:bjab012.

35. Hopper R, Popa D, Maggioni E, et al. Multi-channel portable odor delivery device for self-administered and rapid smell testing. Commun Eng. 2024;3(1):141.

36. Hummel T, Konnerth CG, Rosenheim K, Kobal G. Screening of olfactory function with a four-minute odor identification test: reliability, normative data, and investigations in patients with olfactory loss. Ann Otol Rhinol Laryngol. 2001;110(10):976–81.

37. Coskun V, Ozdenizci B, Ok K. The Survey on Near Field Communication. Sensors (Basel). 2015;15(6):13348–405.

38. Philpott CM, Hummel T, Parma V, Lechner M, Boak D, Obrist M. The Need to Promote Olfactory Health in Public Health Agendas Across the Globe. Clinical Otolaryngology. 2026;51(2):188–93.

