## Supplementary material for "A rapid and connected test for olfactory loss screening"

**Supplementary online material**

**Characteristics of the participants.**

Demographic and olfactory characteristics of the 50 participants are provided in **Table S1.** Mean age was similar between normosmics and dysosmics (W = 354.50, p = 0.42), as was the male-female distribution (*X^2^*_(1)_=0, *p*>0.99). As expected, localization and identification ETOC scores were significantly lower in dysosmics than in normosmics (localization score, W=74.50, p<0.001; identification score, W=0, p<0.001), as was the Sniffin’ Sticks identification score (t_(30)_=-5.10, p<0.001).

**Odor selection process**

The scented cards (manufactured by Euracli®) were based on the “scratch-and-sniff” principle ^(4,39)^, using microcapsule-based ink printed on cards (300g/m2, mat paper; 9cm x 5.8cm). The odorants were deposited on a yellow circular surface of 1.5cm in diameter in the middle of a white card, which was then individually wrapped in plastic packaging. The odors were selected on the basis of a series of technical tests carried out prior to the study. The initial set consisted of 22 odors selected for their compatibility with micro-encapsulation technology (which depends in particular on the solubility of the molecule or mixture used). Of these 22 odors printed on cards, we selected those with a satisfactory perceived quality and intensity. We were left with 13 odors, some of which were subsequently discarded because they were associated with specific anosmia/hyposmia (flower odor of beta-ionone ^(40)^) or because their intensity was not stable over time in a 12-month test (lemon and anise). Two more odors (pine and fuel) were eliminated in a final technical test because of their lower perceived intensity and unstable odor quality. The final set of odorized cards thus included 8 odors mainly from the food sphere (banana, cinnamon, clove, eucalyptus, garlic, mint, orange, thyme, see **Table S2**) whose intensity and perceived quality were stable over time.

**Reference olfactory tests**

*ETOC test*

The ETOC ^(16)^ consists of 16 rows of 4 vials, all filled with odorless mineral oil as a solvent. Each flask contained a synthetic absorbent (polypropylene) to optimize odor diffusion. In each row, only one vial contained an odorant, the others were odorless (solvent alone). The 16 rows correspond to a total of 16 different odorants. Participants had to smell the 4 vials of a row successively and to perform two different tasks. First, participants had to localize the odorized vial among the four. Second, they had to sniff again the vial they selected as being the odorous one, and were asked to identify the odor perceived by choosing between 4 proposed items. A detection/localization score (between 0 and 16) was calculated by adding up the correctly localized odors. Likewise, an identification score (between 0 and 16) was calculated by adding up the correctly identified odors, among the correctly localized ones (in order to limit correct identification by chance). Finally, using these two scores, each participant was classified as normosmic (no olfactory disorder), hyposmic (reduced olfactory perception) or anosmic (complete or almost complete loss of olfaction), according to normative data from a previous study ^(22)^.

*Sniffin’ Sticks identification test*

The Sniffin’ Sticks identification subtest ^(15)^ consists of 16 odorized pens for which the smell must be identified among 4 different proposed labels. Each pen was placed by the experimenter approximately 2cm from the participant’s nostrils, for a duration of 2-3s. A 40s interval was maintained between each pen to minimize olfactory fatigue. A total identification score was calculated from the number of correctly identified odors (between 0 and 16).

**Data analysis**

All statistics were performed using R software ^(23)^. Data normality and homoscedasticity were assessed by Shapiro-Wilk and Bartlett tests. Data are reported as mean±sem or median and first and third quartiles. Alpha value was set to 0.05 for all statistical tests.

A comparison of the normosmic and dysosmic groups in terms of demographic and olfactory characteristics on the reference tests (ETOC, Sniffin’ Sticks) was conducted using Chi-square test for categorical data (sex), and Student t-test or Wilcoxon test for the continuous data (age, olfactory scores) depending on the normality or not of the data, respectively.

To analyze the OS-card test performances, participants being normosmic and dysosmic according to ETOC were compared for four parameters (using Wilcoxon tests, as normality condition was not met, or Chi-Square tests): 1) the intensity ratings (very low=1; low=2; medium=3; strong=4; very strong=5) of each card, 2) the proportion of correct identification of each card (for these two first parameters, Bonferroni corrections for multiple comparisons were applied), 3) a mean perceived intensity score, computed by averaging intensity ratings of the 8 cards for each participant, 4) an identification score set as the number of correct identifications (among the 8 cards) for each participant.

To link the OS-card scores to those of the reference olfactory tests (ETOC, Sniffin’ Sticks), we performed correlation analyses between the perceived intensity score of OS-card and the localization score of ETOC on the one hand, and between the identification score of OS-card and that of both ETOC and the Sniffin’ Sticks on the other hand. Spearman correlation coefficients were used, as normality was not met.

Finally, to evaluate the ability of OS-card to accurately discriminate between the olfactory statuses of the participants (defined with ETOC), we used 3 different linear models on which we performed a discrimination analysis using cross-validation (with a leave one out cross-validation) using the package ‘Caret’ ^(24)^. The main first model aimed to evaluate whether both perceived intensity and identification scores of OS-card could accurately discriminate *normosmics vs dysosmics*. A second model was run to test whether the OS-card identification score alone could be sufficient to discriminate *normosmics vs dysosmics* (as argued in the Sniffin’ Sticks identification subtest procedure; ^(36)^. Last, a third model aimed to assess if OS-card was able to accurately discriminate extreme olfactory statuses, namely *normosmics vs anosmics*. For this last model, both OS-card intensity and identification scores were used. As groups were imbalanced (N=25 for normosmics vs. 11 for anosmics, see **Table S1**), a down-sampling method was used, consisting of removing data from the majority group (normosmics) in order to match the size of the minority group (anosmics). For each of these three models, classification performances were assessed using the Area Under the Curve (AUC) of the mean Receiver Operating Characteristic (ROC), the total correct classification rate (rate of correctly classified participants, all groups considered), the sensitivity (true positive rate, i.e. rate of correctly classified dysosmics or anosmics) and the specificity (false positive rate, i.e. rate of correctly classified normosmics). Misclassified individuals were compared to correctly classified ones in terms of their demographic and olfactory characteristics, with Student t-test or Wilcoxon test depending on the normality of the data. Finally, in all analyses, we used a criterion of probability equal to or higher than 50% to affect a participant to a specific group (normosmics or dysosmics/anosmics).

**References**

1.	Manesse C, Ferdenzi C, Sabri M, et al. Dysosmia-associated changes in eating behavior. Chemosensory Perception. 2017;10(4):104–13.

2.	Manesse C, Ferdenzi C, Mantel M, et al. The prevalence of olfactory deficits and their effects on eating behavior from childhood to old age: A large-scale study in the French population. Food Quality and Preference. 2021;93:104273.

3.	Razafindrazaka H, Pereda-Loth V, Ferdenzi C, et al. African Gene Flow Reduces Beta-Ionone Anosmia/Hyposmia Prevalence in Admixed Malagasy Populations. Brain Sci. 2021;11(11):1405.

4.	Thomas-Danguin T, Rouby C, Sicard G, et al. Development of the ETOC: a European test of olfactory capabilities. Rhinology. 2003;41(3):142–51.

5.	Joussain P, Bessy M, Faure F, et al. Application of the European Test of Olfactory Capabilities in patients with olfactory impairment. Eur Arch Otorhinolaryngol. 2016;273(2):381–90.

6.	Hummel T, Sekinger B, Wolf SR, Pauli E, Kobal G. ‘Sniffin’ Sticks’: Olfactory Performance Assessed by the Combined Testing of Odour Identification, Odor Discrimination and Olfactory Threshold. Chem Senses. 1997;22(1):39–52.

7.	R Core Team. R: A Language and Environment for Statistical Computing. 2023; Available from: https://www.R-project.org/

8.	Kuhn M. Building Predictive Models in R Using the caret Package. Journal of Statistical Software. 2008;28:1–26.

9.	Hummel T, Konnerth CG, Rosenheim K, Kobal G. Screening of olfactory function with a four-minute odor identification test: reliability, normative data, and investigations in patients with olfactory loss. Ann Otol Rhinol Laryngol. 2001;110(10):976–81.
