## Supplementary material for "A rapid and connected test for olfactory loss screening": Table S1

**Table S1.** Participants were classified as normosmic or dysosmic based on their ETOC scores. Data are represented as mean ± sem. n = sample size.

|  | **Normosmics** | **Dysosmics** | **Test** | **p** |
| --- | --- | --- | --- | --- |
| **N** | 25 | 25 |  | |
| **Age (years)** | 60.7 ± 2.47 | 62.9 ± 2.39 | Wilcoxon | 0.42 |
| **Women (n)** | 16 | 17 | Chi-Square | >0.99 |
| **Men (n)** | 9 | 8 |  |  |
| **ETOC localization score** | 15.7 ± 0.09 | 10.2 ± 1.01 | Wilcoxon | **<0.001** |
| **ETOC identification score** | 13.5 ± 0.25 | 5.2 ± 0.77 | Wilcoxon | **<0.001** |
| **Sniffin’ Sticks identification score** | 12.9 ± 0.30 | 8.4 ± 0.83 | Student t-test | **<0.001** |
| **Hyposmics (n)** |  | 14 |  | |
| **Anosmics (n)** |  | 11 |  |  |
| **Disorder duration (years)** |  | 7.1 ± 2.61 |  |  |
