## Supplementary material for "A rapid and connected test for olfactory loss screening": Table S2

**Table S2.** List of odors used in the OS-card test, including their quality, compositions when available, provider, ID (Compound Identifier: CID or reference), concentrations and labels provided in the identification task.

|  | **Simple compounds** | | | | **Complex aromas** | | | |
| --- | --- | --- | --- | --- | --- | --- | --- | --- |
| **Quality** | Banana | Clove | Eucalyptus | Mint | Cinnamon | Garlic | Orange | Thyme |
| **Composition (provider)** | Isoamyl acetate  (Sigma-Aldrich) | Eugenol  (Sigma-Aldrich) | Cineol  (Sigma Aldrich) | L-carvone  (Sigma-Aldrich) | Aroma (Euracli®) | Aroma (Euracli®) | Aroma (Euracli®) | Aroma (Euracli®) |
| **ID (CID number or reference)** | 31276 | 3314 | 2758 | 439570 | FE07627 | FE02314 | FE05480 | FE02202 |
| **Mass concentration (%)** | 20.20 | 21.50 | 21.50 | 21.50 | 21.30 | 22 | 20.80 | 21.80 |
| **Labels options (identification)** | Almond  Lemon  Carrot  Banana | Lavender  Clove  Chives  Mushroom | Violet  Strawberry  Eucalyptus  Tomato | Nutmeg  Olive  Basil  Mint | Cinnamon  Hazelnut  Butter  Coffee | Cabbage  Garlic  Leek  Celery | Pear  Banana  Orange  Blackberry | Thyme  Licorice  Parsley  Pepper |
